# Closing the gap towards a successful referral system. A case study of a tertiary teaching and referral hospital, Kenya: pre-posttest study design

**DOI:** 10.1101/2024.01.02.24300732

**Authors:** Maxwell Philip Omondi

## Abstract

Inappropriate utilization of higher-level health facilities and ineffective management of the referral processes in resource-limited settings is increasingly becoming a concern in health care management in developing countries. This is characterized by self-referrals and frequent bypassing of nearest health facilities. On 1^st^ July 2021, Kenyatta National Hospital (KNH) did enforce the national referral guidelines that required patients have a formal referral letter to reduce the number of self-referrals, decongest KNH and allow KNH to function as a referral facility as envisioned by Kenya Health Sector Referral Implementation Guidelines of 2014, Kenya 2010 constitution and KNH legal statue of 1987. The purpose of this study was to analyse the effect of enforcement of referral guidelines on facility referrals to KNH.This was a pre-posttest study design. The study was conducted amongst the orthopedic facility referrals in 2021 with 222 and 246 before and after enforcement of referral guidelines respectively. Data collection was done through data abstraction. Data was analyzed using frequency distribution, pearson chi-square test and logistic regression. Nairobi County and its environs constituted over four-fifth of all facility referrals to KNH. Over two-thirds of the facility referrals to KNH were from government facilities. There was significant reduction in health facilities tiers 2 and 3 referring patients directly to KNH after enforcement of referral guidelines (p=0.002). About 43 health facilities ceased referring patients to KNH with over two-thirds of these health facilities being private facilities. The major facility and patient factors that were associated with facility referrals to KNH were human resource capacity and availability and patient’s preference. In conclusion, enforcement of the referral guidelines significantly reduced the lower tiers health facilities referring to KNH. We recommend having written standard operating procedures on referrals based on the national referral guidelines with continued enforcement of the same to sustain the gains made.

## Background

Tertiary hospitals in resource-limited countries treat patients referred but in most cases are the first level of care for the vast majority of patients (1). One of the challenges in health care delivery in resource-limited settings is inappropriate utilization of tertiary health facilities that results in patients’ congestion in referral hospitals with simple medical conditions that can be effectively managed at the lower peripheral health facilities. The majority of these patients are self-referred, bypassing lower-level health facilities in the process (2–5).

A study done in Lusaka demonstrated how University Teaching and Referral Hospital is congested due to bypassing of lower peripheral health facilities and as a result the tertiary facility effectively functions as a primary health care facility. The urban phenomenon of widespread self-referral is associated with low rates of formal referral from peripheral health facilities (4, 6). The net result is simple medical conditions end up being managed in high-cost referral health facilities leading to overcrowding, long waiting times, and scarce staff time consumed by lesser medical conditions at the expense of complex medical conditions. It is therefore imperative that attempts need to be made to ensure patients make use of lower health facilities by creating disincentives for patients bye-passing these health facilities (1, 7). The bypassing of lower level health facilities also cripples the primary health care system and this effectively ensures that primary health care facilities remain underused and inefficient (8). Taken together, this jeopardizes the appropriate delivery of primary, secondary, and tertiary health care

In 2008, an assessment done in South African Public health sector revealed that patients were accessing the health system at inappropriate levels and bypassing primary health care to attend to regional and tertiary hospitals as their initial visit leading to overcrowding and unnecessary costs to the referral facilities (9). In cross-sectional study was done to review the self-referrals to a District-Regional Hospital in KwaZulu Natal, South Africa showed 36% were self-referrals. However, majority (64%) were appropriately referred with written referral letters (9).

A study on referral system in Nigeria showed that 92.9% reported to tertiary hospital directly without referral while 7.1% were referred. The result of this is overcrowding of the tertiary facilities with conditions that can be managed at lower level health facilities (7).

In India health care system is characterised by overcrowding, lack of specialist doctors and effective referral system. Despite having a referral guideline that details referral of patients from lower levels to sub-district, district and then to tertiary hospitals, the actual practices are that patients go to any level of health care system without any referral (10). This creates burden on the secondary and tertiary level hospitals. In Taiwan where we have a free-access systems, bypass of primary care and seeking secondary care through self-referral is rampant despite Taiwan’s government taking various initiatives to mitigate bypass (11).

Kenya is a low-income middle-income country. Kenya’s health sector is one of the 14 functions devolved to the 47 county governments as provided for in the Fourth Schedule of the 2010 constitution (12). There are six different levels or tiers of health care in Kenya. Tiers 1 -5 are managed by the county government and tier 6 by the national government. In this system patients are required to move from one tier to the next using a formal referral letter. Kenya has a pluralistic health sector consisting of private and non-governmental service providers alongside the government-run health facilities that is organized into six (6) tiers (Figure 1).

**Figure 1:**
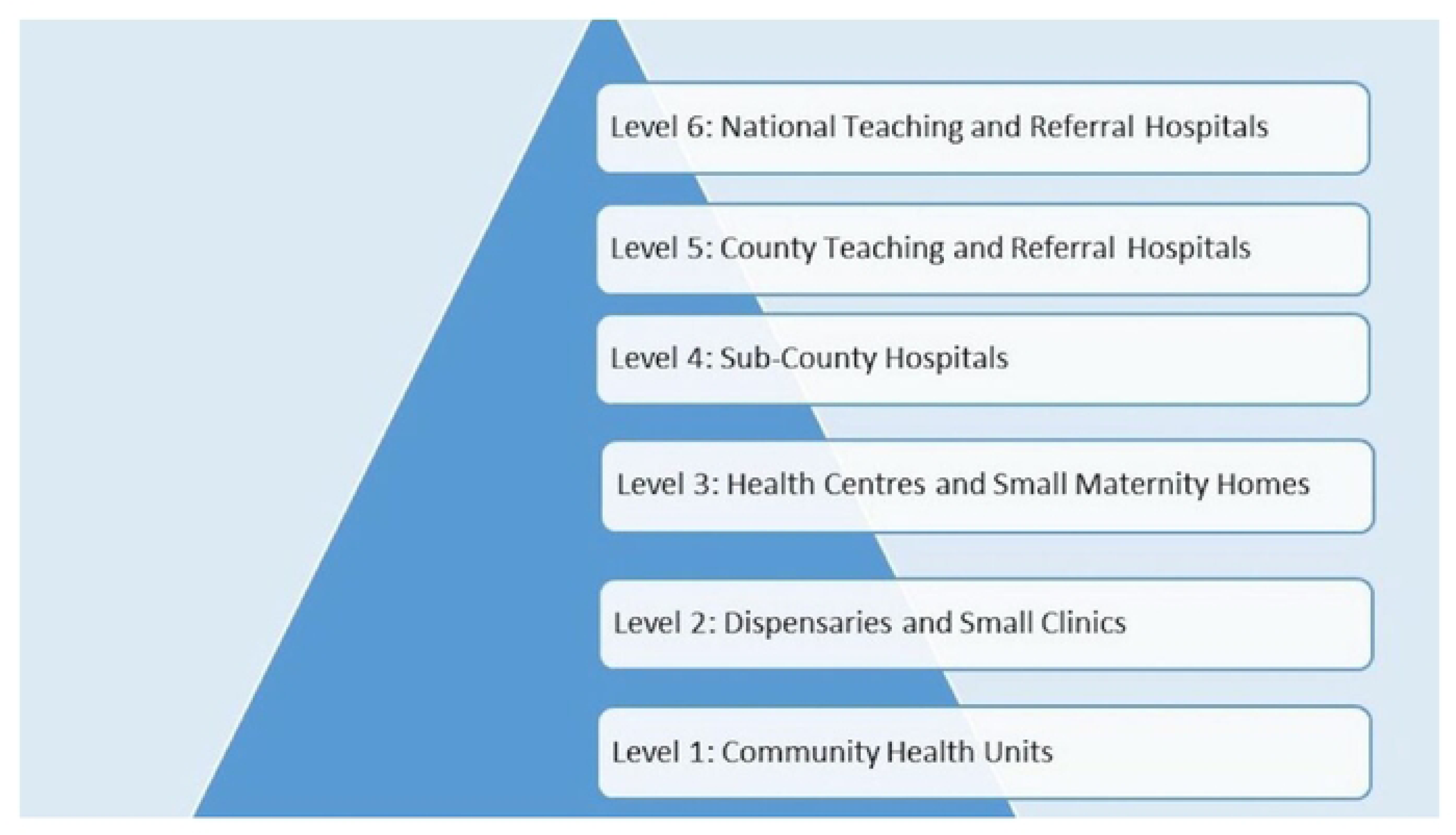
Six tiers of health care service delivery in Kenya (13)

Tiers 4 are Sub-County Hospitals, Nursing Homes and also include medium-sized private hospitals. Tier 5 are County Teaching and Referral Hospitals, large private as well as faith-based hospitals. Tier 6 are National Teaching and Referral Hospitals. In Kenya there are four teaching and referral hospitals: a) Mathari Hospital, b) Kenyatta National Hospital, c) Moi Teaching and Referral Hospital and d) National Spinal Injury Hospital. These tier 6 offer range of services similar to Tier V but additionally they offer specialised treatments in addition to having advanced infrastructure and are not only accessed by Kenyans but so serve East Africa and Central Africa.

Kenyatta National Hospital (KNH) was established as a National Referral and Teaching Hospital, to provide training and medical research. KNH was established in 1901 and became a State Corporation in 1987 and sits at the peak of the health referral system in Kenya (14). According to the KNH Board order of 1987 contained in the Legal Notice No. 109, the functions of KNH were spelled out as a) to receive patients on referral for specialized health care; b) to provide facilities for medical education for the University of Nairobi and other health allied courses; c) to contribute to national health planning (14). This understanding has been reinforced by the Kenya Health Sector Referral Implementation Guidelines, 2014, and the Constitution of Kenya 2010 which tasks KNH with the responsibility for health policy formulation (12, 15).

Orthopaedic wards in KNH have consistently recorded the highest bed occupancy percent for the last couple of years. In 2018, 2019 and 2020 it recorded bed occupancy percent of 142.2%, 138.2% and 116.5% respectively against the KNH bed occupancy percent of 106.2%, 113.4% and 91.5% (16). This coupled with low nurse-patient ratio of 1:10 compromises not only the quality of nursing care given to patients but also the ability of KNH to effectively perform its statutory obligations. Various studies have demonstrated that an appropriate nurse-patient ratio is associated with the reduction in medical errors, decubitus ulcers, hospital-acquired infections, long duration of hospital stay, high readmission rates, and compromised patient well-being and safety due to the nurse burnout (17–19).

On 1^st^ July 2021, KNH management made a decision to enforce the Kenya Health Sector Referral Guidelines 2014 that places KNH at the tip of the health sector referral system. This meant that patients will be seen based on referral letters from other health facilities to reduce the number of walk-in patients who would have otherwise been appropriately seen at the peripheral health facilities. This would then allow KNH to focus on the management of complex medical conditions and allow uncomplicated medical conditions to be managed at lower-level health facilities. The purpose of this study was to analyse the effect of enforcement of referral guidelines on facility referrals to KNH. The findings of this study will help formulate policy and guidelines on orthopaedic and trauma essential care at the peripheral and tertiary health facilities in Nairobi Metropolitan Area and the country at large. It will also provide evidence-based information that will contribute to the review of Kenya Health Sector Referral Implementation Guidelines of 2014.

## Methods

### Study design

This was a pre-posttest study design. The national referral guideline was enforced on 1^st^ July, 2021. The pretest covered 5 months before enforcement of referral guidelines (February 1, to June 30, 2021) while post-test covered 5 months after enforcement of the referral guidelines (August 1, to December 31, 2021). The variables compared were county, type of health facility (government or private) and health facility tier before and after enforcement of the referral guidelines. Enforcement of referral guidelines required that the referring health facility consults with KNH referral Office for concurrence before patients are referred to KNH and that patients should come with written official referral letters. This was to ensure only patients who require specialized orthopaedic and trauma care not available at the peripheral health facilities get admitted to KNH.

### Study area

KNH is the largest teaching and referral hospital in East and Central Africa. KNH Orthopaedic Wards were the study area. KNH is based in Upperhill, Nairobi, the capital city of Kenya. It is located along Hospital Road, about 5km from the city centre. KNH has a bed capacity of 1,800, 6,000+ staff members, 50 wards, 22 out-patient clinics, 24 theaters (16 specialized) and Accident & Emergency Department (14). Of the 1800 bed capacity, 96 beds are allocated to orthopaedic wards. KNH is a 10-floor storied building complex and the Orthopaedic wards are located on the 6^th^ floor but we also have orthopaedic admissions in private wings on 9^th^ and 10^th^ floor. Orthopaedic patients with other co-morbidities also get admitted to other wards in KNH.

### Study duration

The study duration was from 1^st^ February 2021 to 31st December 2021. The referral guidelines were enforced from 1^st^ July 2021. Data abstraction was conducted from 1^st^ January to 31^st^ March, 2022.

### Study population

Orthopaedic inpatient caseload before and after enforcement of referral guidelines.

### Eligibility criteria

#### Inclusion criteria

All orthopaedic and trauma facility referrals to KNH during the study period.

### Sample size calculation

Sample size was calculated using an adjusted Casagrande formula for calculating sample sizes that compare two binomial distributions (20).

A sample size of 468 facility referrals were enrolled during the study period with 220 and 248 facility referrals before and after the enforcement of national referral guidelines.

### Recruitment and sampling procedures

Three (3) research assistants (RAs) were recruited to collect and abstract patient data from patient files. The RAs were health care workers with a diploma in Orthopedic Trauma and with some experience in research data collection. The Principal Investigator (PI) was the research coordinator for the data collection. The orthopedic and trauma admissions with facility referrals were identified from the a) admission desk of Health Information System at KNH Accident and Emergency Unit (A&E) b) KNH Orthopedic Outpatient clinic records (OC) c) KNH Corporate Outpatient Care (COC). They were then recorded in a logbook. This logbook served as a master register for all facility referred patients admitted and therefore the sampling frame for the study. All facility admissions were logged into the logbook from the admission books stationed in these three (3) services points. Proportional Population to Size (PPS) was then used to decide on the numbers to be sampled per month from each of these three services points so that the sample size would be a representative of the admissions by month from each of these three orthopedic admissions entry points (Table 1).

**Table 1:**
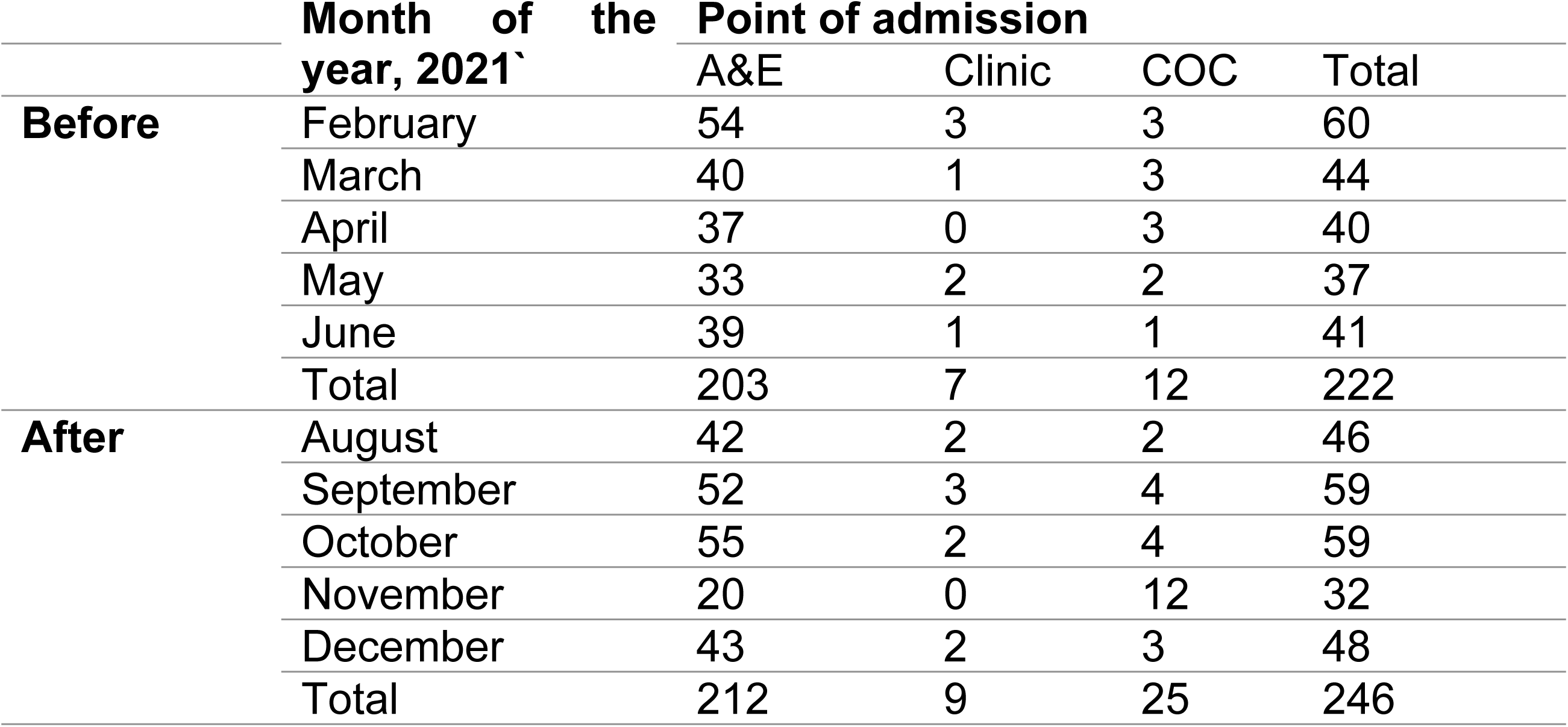
Orthopaedic and trauma admissions to KNH stratified by point of admission, 2021.

The three (3) RAs were reporting to and working under the direction of the PI. The RAs were trained for two (2) days by the PI on the research protocol, data collection tools, data collection procedures and that included pilot testing of the data collection tools as well before the actual data abstraction.

A written Informed consent was obtained from KNH Medical Research Department to have access to the patient’s health records in the Health Information Office (Room 19).

### Variables

a. Name of health facility County c) Facility Type (Government/Private) d) Health Facility Level/Tier e) Admission date (dd/mm/yyyy)

### Data collection procedures

Data collection was done through a data abstraction form from the patient files.

#### Data Abstraction form

The three (3) RAs were trained on the data abstraction using a data abstraction form as per the research protocol. The PI reviewed all the filled-in abstraction forms for completeness and accuracy daily during the entire data collection period and providing regular feedback to the RAs on a timely manner to ensure data quality and compliance to the study protocol. All the completed and verified data abstraction forms were then collected and filed by the PI at the end of every week under a lockable cabinet.

### Ethical considerations

The study protocol was presented to UoN/KNH Ethics and Research Committee and was granted ethical approval (ERC No: P852/10/2021). Administrative approval was also granted by KNH Medical Research Department and KNH Orthopaedics Department.

### Data management, analysis, and presentation plan

Data abstraction tool was designed to collect quantitative and qualitative data. For anonymity and confidentiality purposes the data abstraction tool were marked only with the participant study numbers and no names were used. However, the research assistants and the PI had access to the patients charts and therefore could identify individual patients during the data collection period. The data were entered into a password-protected Redcap database kept by the KNH Medical Research Department. The data were analyzed using SPSS version 21.0. Descriptive statistics such as frequencies while inferential statistics using Pearson’s chi-squared tests, logistic regression were used. The calculations were done at a 95% level of confidence.

## Findings

### Basic Profile of the sample population

A total of 468 charts were abstracted of which 220 (47%) were before and 248 (53%) were after the enforcement of the referral guidelines.

Nairobi County constituted more than half of facility referrals to KNH. This rose to over four – fifths of facility referrals with inclusion of the neighbouring counties namely Kajiado, Kiambu and Machakos. Almost all the facility referrals were from within Kenya (Table 2).

**Table 2:**
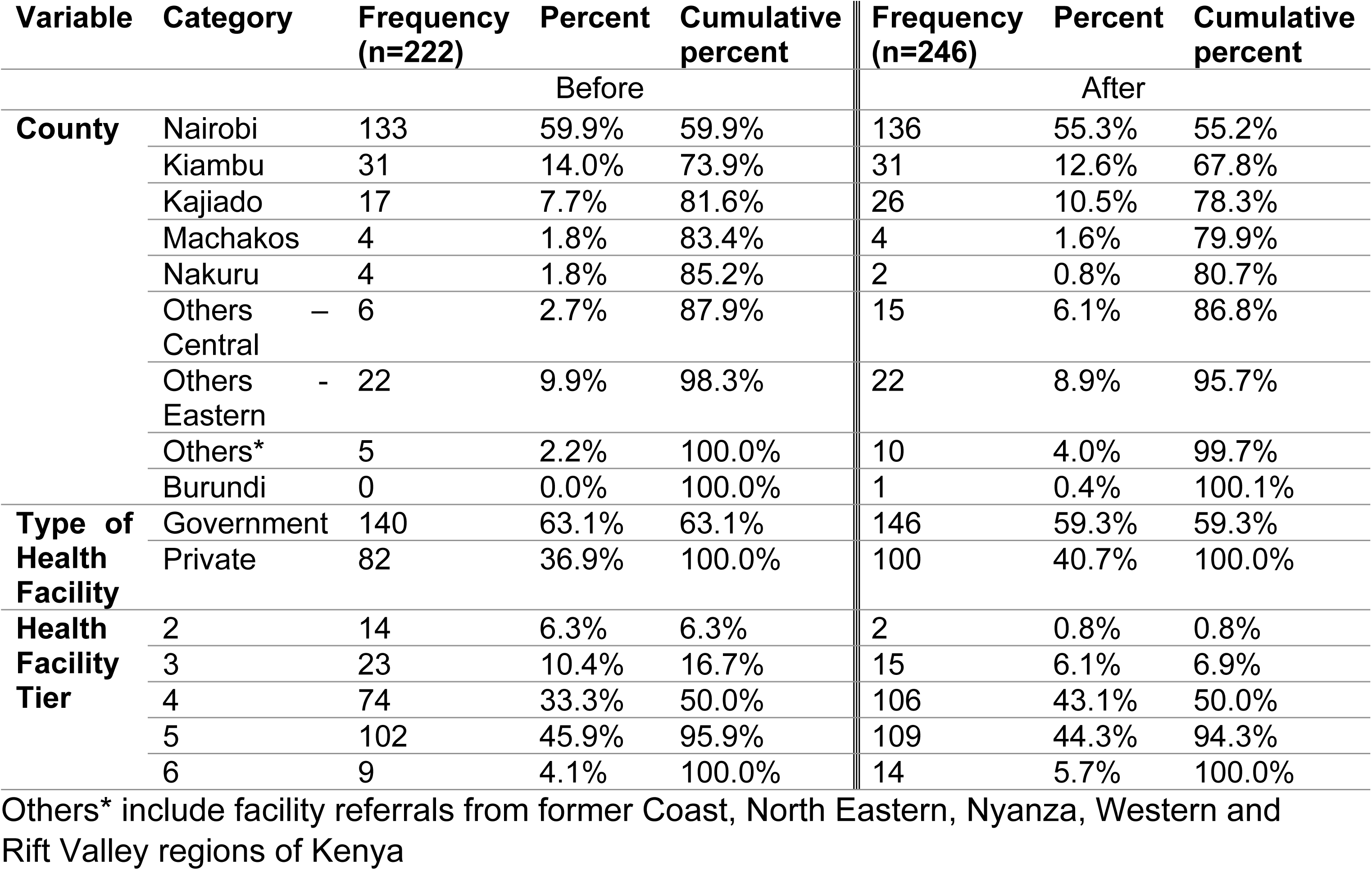
Basic profile of the sample population, 2021.

About three-fifths of the facility referrals to KNH were government health facilities with private health facilities comprising about two-fifths. About half of the facility referrals are from Tiers 2 to 4 and with the remainder of facility referrals to KNH from Tier 5 and 6 (Table 2).

Mama Lucy Kibaki Hospital, Mbagathi District Hospital and Thika Level 5 Hospital were the major health facilities referring patients to KNH before and after enforcement of the referral guidelines (Figure 2).

**Figure 2:**
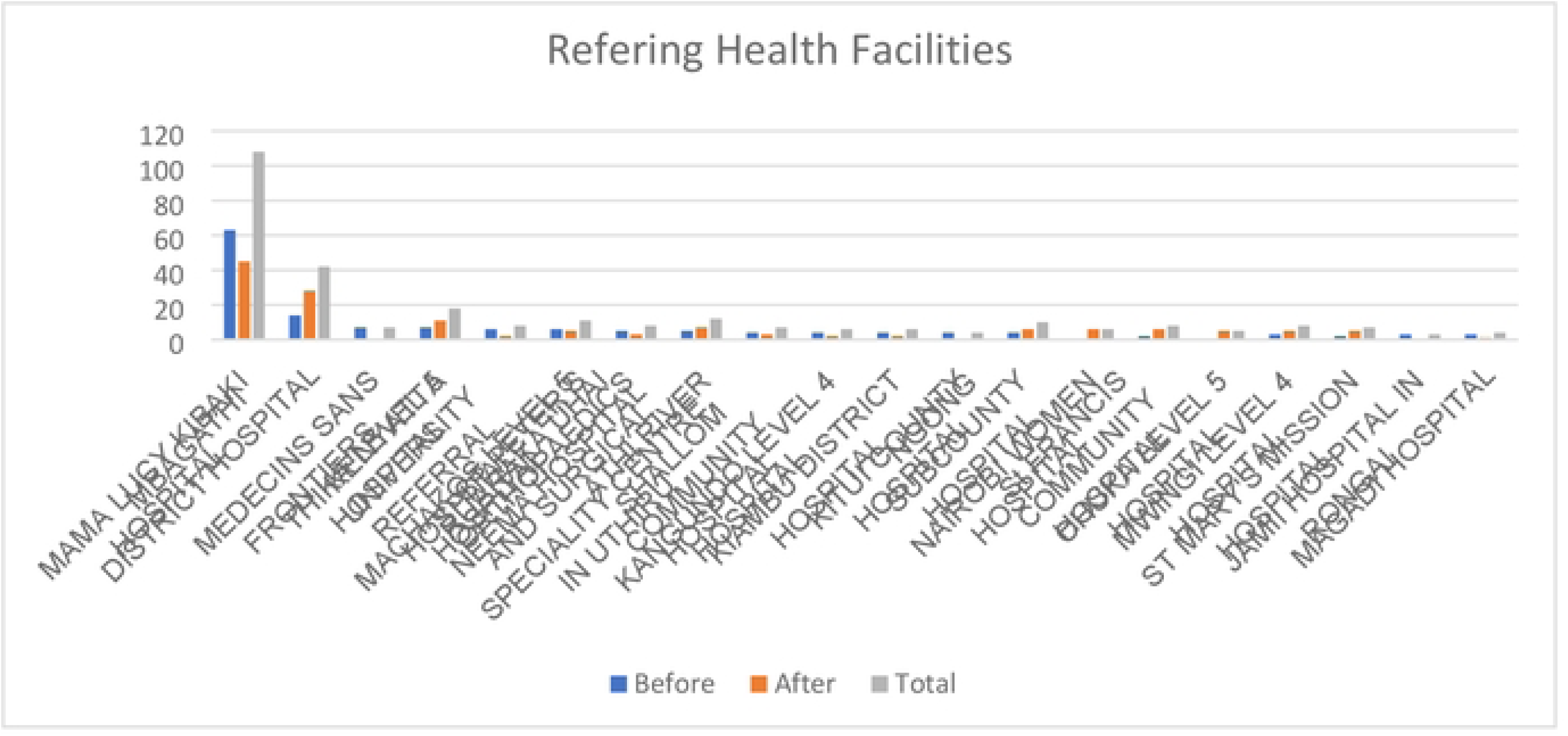
The major health facilities referring to KNH before and after enforcement of referral guidelines, 2021

There were increases in the health facility referrals to KNH after the enforcement of the referral guidelines. However, these increases were not statistically significant differences (Table 3).

**Table 3:** Bivariable analysis of the type of health facilities referrals referring to KNH before and after enforcement of referral guidelines, 2021.

|  |  | Before<br>(n=222) | After (n=246) | p-value |
| --- | --- | --- | --- | --- |
| **Type of Health Facility** | Government Facility referrals | 140 (49.0%) | 146 (51.0%) | p= 0.411 |
|  | Private Facility referrals | 82 (45.1%) | 100 (54.9%) |  |

Legend: Pearson $\chi^2$ statistics test was used to test for statistical significance at 95% CI.

There were statistically significant differences in health facilities referrals to KNH before and after the enforcement of the referral guidelines when stratified by the health facility tier (Table 4). There was significant reduction in Tiers II and III referring patients directly to KNH and an increase in the facility referrals to KNH from tiers IV, V and VI (Table 4). There were less tier II and III health facilities referring patients to KNH after enforcement of the referral guidelines. The facility referrals from Tier II were 90.8% less likely as compared to Tier III after enforcement of the referral guidelines (p=0.006) (Table 4).

**Table 4:** Multivariable analysis of the health facilities referrals, stratified by health facility tier, referring to KNH before and after enforcement of referral guidelines, 2021.

|  |  | Before<br>(n=222) | After (n=246) | Chi-square; p-value | Logistic regression |
| --- | --- | --- | --- | --- | --- |
| **Health Facility Tier** | II | 14 (87.5%) | 2 (12.5%) | 16.505;<br>p=0.002 | 1.0 |
|  | III | 23 (60.5%) | 15 (39.5%) |  | 0.092 (p=0.006) |
|  | IV | 74 (41.1%) | 106 (58.9%) |  | 0.419 (p=0.108) |
|  | V | 102 (48.3%) | 109 (51.7%) |  | 0.921 (p=0.856) |
|  | VI | 9 (39.1%) | 14 (60.9%) |  | 0.687 (p=0.301) |

Legend: Pearson $\chi^2$ statistics test was used to test for statistical significance at 95% CI and Odds Ratio was used to test the strength of association. 1.0 refers to reference group.

While the study showed about 43 health facilities ceased referring patients to KNH with over two-thirds of these health facilities being private facilities (Table 5).

**Table 5:** Frequency distribution of type of health facilities that stopped referring, stratified by Type of Health Facility and Facility Tier, after enforcement of the referral guidelines, 2021.

|  | Categories | Frequency<br>(n=43) | Percent | Cumulative<br>percent |
| --- | --- | --- | --- | --- |
| **Type of Health Facility** | Government | 14 | 32.6% | 32.6% |
|  | Private | 29 | 67.4% | 100.0% |
| **Facility Tier** | II | 7 | 16.3% | 16.3% |
|  | III | 12 | 27.9% | 44.2% |
|  | IV | 16 | 37.2% | 81.4% |
|  | V | 6 | 14.0% | 95.4% |
|  | VI | 2 | 4.7% | 100.1% |

The study revealed that lower tiers health facilities namely tier II-IV ceased referring patients to KNH and this comprised over four-fifths of all health facilities that stopped referring to KNH after enforcement of the referral guidelines (Table 5).

The major facility and patient factors that were associated with facility referrals to KNH were human resource capacity and availability, health facility infrastructure, Orthopaedic equipment’s and implants availability, patient’s preference, financial considerations (Table 6). It’s worth noting that across both private and government health facilities the main factors for referral were inadequate human resource capacity and availability followed by availability of medical supplies and equipments and financial considerations (Table 6). Amongst government health facility referrals, whereas financial considerations was among the common factors for facility referrals to KNH, the patients actually were opting to be referred to KNH as opposed to a private health facility due to cost “…*the patient needed a total knee replacement which could not be done in Nyahururu and they did not have enough funds to take him to a private hospital so they preferred to KNH…*” *‘…. the private hospital was too expensive so they opted for KNH…’*

**Table 6:**
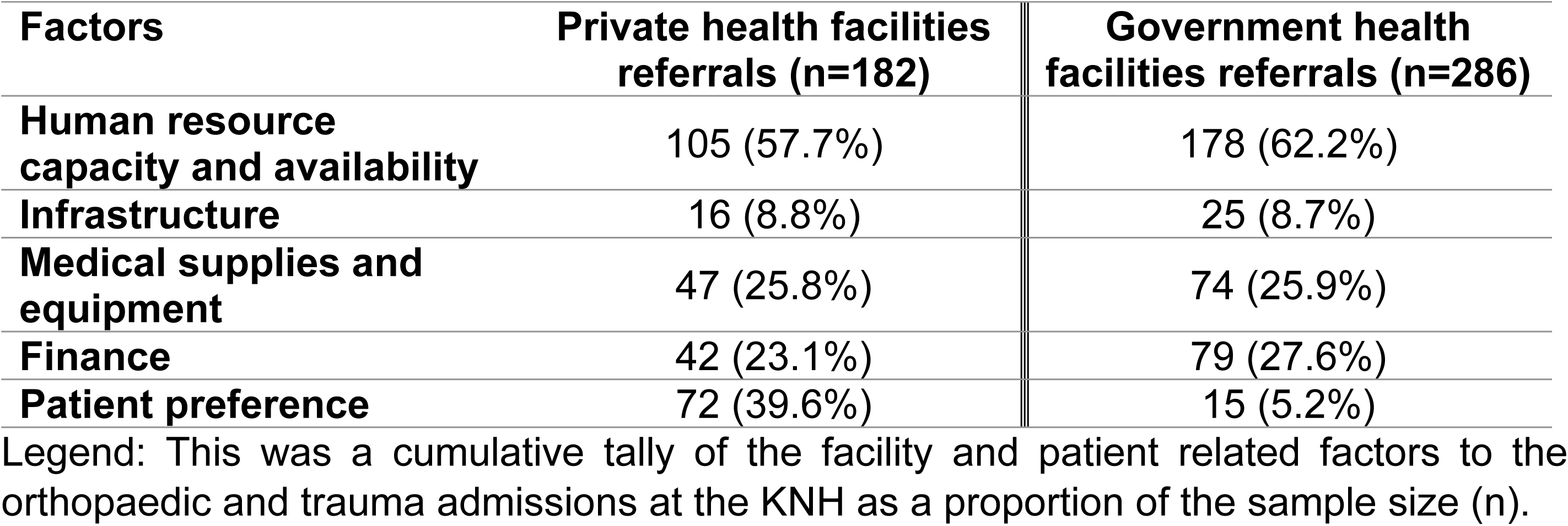
Table showing the frequency distribution of the major facility and patient related factors to the orthopaedic and trauma admissions at the KNH.

| Factors | Private health facilities referrals (n=182) | Government health facilities referrals (n=286) |
| --- | --- | --- |
| Human resource capacity and availability | 105 (57.7%) | 178 (62.2%) |
| Infrastructure | 16 (8.8%) | 25 (8.7%) |
| Medical supplies and equipment | 47 (25.8%) | 74 (25.9%) |
| Finance | 42 (23.1%) | 79 (27.6%) |
| Patient preference | 72 (39.6%) | 15 (5.2%) |

Legend: This was a cumulative tally of the facility and patient related factors to the orthopaedic and trauma admissions at the KNH as a proportion of the sample size (n).

Patients’ preference was shown to be a significant factor associated with private health facility referrals to KNH as opposed to government health facility referrals to KNH (Table 6).

## Discussions

While orthopaedic facility referrals were spread across the country, over four – fifths of these facility referrals were from Nairobi County and its environs. This is in tandem with studies done at tertiary teaching and referral health facilities in Rwanda, Tanzania, Malawi and Nigeria that revealed significant majority of the facility referrals are from within the region where the health facilities are co-located (7, 21–24).

Most of the facility referrals to KNH are from government health facilities with private health facilities comprising about one-third. This compares favourably with a study done in Rwanda that showed approximately 85.7% of the facility referrals were from public hospitals while 10.7% of the referrals were from private health facilities (25). Similarly a prospective study done at Muhimbili National Hospital, Tanzania to examine the medical referral pattern of patients in 2004, revealed that about 22.4% of admissions were from public health facilities while only 4.2% were from private health facilities (26).

Mama Lucy Kibaki Hospital, Mbagathi District Hospital and Thika Level 5 Hospital were the major health facilities referring patients to KNH before and after enforcement of the referral guidelines and they all happen to be government health facilities – which comprise majority of health facility referrals to KNH as compared to private health facility referrals. This is probably because of inadequate human resource, inadequate infrastructure like bed capacity, theatre space as well as lack of orthopaedic equipments and implants. However, this contrasts with a study on referrals to a tertiary health facility in Nigeria that showed over half of the admissions were from private health facilities (7). In addition, St Peters Orthopaedic and Surgical Speciality Centre, St Francis Community Hospital, Arthi River Shalom Community Hospital and St Mary’s Mission Hospital were the most frequently referring private health facilities to KNH for orthopaedic admissions in 2021. These were probably due to inadequate human capacity, patients’ preference and financial considerations.

There were increases in the number of health facilities referring patients to KNH after the enforcement of the referral guidelines. This is in tandem with a study done in Republic of Hondura that showed the referral rate was observed to be higher when institutional managers emphasized the importance of the referral system (4). The enforcement of the national referral guidelines meant more awareness on the referral requirements and importance of referrals of patients to the next level of care and this might have resulted in increase in the number of health facilities referring patients.

The enforcement of the national referral guidelines was meant to streamline the referral process from the peripheral health facilities to KNH and in the process allow KNH to manage complex orthopaedic cases that cannot be handled at the lower-level health facilities. All potential referring health facilities were required to refer patients to the next tier of health facility instead of referring directly to KNH and the higher tier health facilities were expected to seek concurrence before referring patients to KNH and that should be accompanied with an official referring letter. Due to the enforcement of these referral requirements, there was significant reduction in Tiers 2 and 3 referring patients directly to KNH and an increase in the patients being referred to KNH from tiers 4, 5 and 6.

The study showed about 43 health facilities ceased referring patients to directly to KNH with over two-thirds of these health facilities being private facilities. This is in tandem with a study on Geographic accessibility to public and private health facilities done in Kenya in 2021 that revealed the private health facilities’ distribution was skewed toward the urban counties (27). The implication of this is that a good number of private health facilities were no longer referred patients directly to KNH.

Lower tiers health facilities namely tier 2 and 3 largely reduced patient referrals to KNH and this together with tier 4 comprised over four-fifths of all health facilities that stopped referring to KNH after enforcement of the referral guidelines. This is because these lower-tier facilities were now required to refer patients to the nearest Tier 4 and 5 health facilities as their first referral point. This means that the enforcement of the national referral guidelines did reduce referrals from lower tier health facilities and probably were referring them to the next tier facility. This may explain why the numbers of higher tiers referring patients to KNH increased after the enforcement of the referral guidelines.

Human resource capacity remains a major factor in health facility referrals both for private and government health facilities. Inadequate human resource does also comprise the quality of care provided at the health facilities. This is in tandem with a study finding in Kenya that revealed Human resources for health was inadequately financed with insufficient number of health workers and there was maldistribution of staff in favor of higher-level facilities and in urban cities and this resulted in unnecessary referrals to higher level consequently compromised quality of primary healthcare (28–30). A number of studies in Brazil, Japan, United States of America and in Low- and Middle-income countries have also demonstrated inadequate human resource capacity and availability as the main reasons for patient referrals (31–35). However, this contradicts a study done in China on types of health care facilities and the quality of primary care revealed that Community Health Centers offer quality health care services than the secondary and tertiary health care hospitals (36). This was largely due to the fact that it was Chinese deliberate government policy to support and improve the capacity of the of the Community Health Centers.

Private health facility referrals to KNH were to significant extent dictated by patients’ preference. This could be due to perceived good quality of care but also due to perceived low cost of medical care in KNH by virtue of it being a government national referral hospital. Studies done in California, USA and Islamic Republic of Iran, Cape Town, South Africa did show that the perception of good quality of care does influence choice of health facility (3, 37–39).

The study had a few limitations. First, the effect of COVID 19 pandemic on facility referrals of cases from peripheral health facilities and walk-in patients. This was mitigated by ensuring the data collection period covered the covid period where intercounty movement restrictions were lifted. secondly, this study is retrospective and a quasi-experimental study design and hence weaker in determining causality. Despite these limitations, given the paucity of published literature in this study topic, this study offers key information on the effects of enforcement of referral guidelines in health facility referrals to KNH with important lessons for Kenya and possible sub-Saharan Africa.

## Conclusions

The enforcement of the national referral guidelines did reduce the number of facility referrals from lower tier health facilities with a good number of private health facilities stopped referring patients to KNH altogether. The major facility and patient factors that were associated with facility referrals to KNH were human resource capacity and availability, health facility infrastructure, Orthopaedic equipment’s and implants availability, patient’s preference, financial considerations. Therefore, it is possible to successfully enforce the implementation of referral guidelines.

## Recommendations

### Recommendations to Policy Makers/ County Government

1. Regularly sensitize the health facilities on the implementation of the national referral guidelines 2014 for continued enforcement of the referral guidelines;
2. Strengthen the human resource capacity and availability to minimize on referrals to KNH and this will consequently strengthen the primary and tertiary level orthopaedic and trauma care and help decongest KNH and allow it to function as a premier referral facility as per its statutory obligations;
3. Develop, disseminate and train health facilities on standard operating procedures based on the national referral guidelines and ensure adherence to the same.

### Recommendations to KNH

1. Educate and sensitize the health facilities on the role of KNH as a premier National Teaching and Referral facility that is mandated to manage complex referrals and should not be the first point of contact for patients seeking orthopaedic care;
2. Develop, disseminate and train KNH staff from A&E department, Orthopedic Clinic and Corporate Outpatient Centre on the national referral guidelines and referral standard operating procedures.

### Recommendations to Health Facilities

1. Ensure adherence to the national referral guidelines of 2014 for an effective referral process and to ensure optimum utilization of primary, secondary and tertiary level of care.

## Data availability

Data is available and has been uploaded in the submission portal.

## Conflict of interests

The author has no conflict of interest to declare.

## Funding statements

This study was partially funded by Kenyatta National Hospital RFA 2020/21.

## Acknowledgements

We would like to sincerely acknowledge the work of Brian Okinyi and Micah J. Kipkemei for commitment and assistance in data collection process.

## Captions of supporting Information files

S1 Supporting Information – Working dataset 04.12.2023

S2 Supporting Information – Facilities stopped referring

S3 Supporting Information – Reasons for Referral spreadsheet

S4 Supporting Information – Reasons for Referral Tally spreadsheet

S5 Supporting Information – Referrals spreadsheet

S6 Supporting Information – Referral facilities spreadsheet

Other - ERB approval letter - ERC APPROVAL LETTER.pdf

Other - PLOSOne_Human_Subjects_Research_Checklist.doc

## Notes

### Competing Interest Statement

The authors have declared no competing interest.

### Funding Statement

This study was partially funded by Kenyatta National Hospital. RFA 2020/21. The funders had no role in study design, data collection and analysis, decision to publish and in preparation of the manuscript.

### Author Declarations

University of Nairobi/KNH Ethical Review Board. The study protocol was presented to UoN/KNH Ethics and Research Committee and was granted ethical approval (ERC No: P852/10/2021).

